# Clinical usage of non-invasive ventilation by physical therapists in cystic fibrosis centres across Australia: A cross-sectional survey study

**DOI:** 10.1101/2021.06.30.21259429

**Authors:** Molly O Foxcroft, Rebecca Chambers, Robyn Cobb, Suzanne Kuys, Kathleen Hall

**Affiliations:** School of Allied Health, Faculty of Health Sciences, Australian Catholic University, 1100 Nudgee Road, Banyo, QLD, 4014, Australia; Physiotherapy, Adult Cystic Fibrosis Centre, The Prince Charles Hospital, 627 Rode Road, Chermside, QLD, 4032, Australia

**Keywords:** Cystic Fibrosis, Noninvasive Ventilation, Bilevel Positive Airway Pressure, Physical Therapy, Airway Clearance, Exercise

## Abstract

**BACKGROUND:** This study investigated clinical usage of non-invasive ventilation during physical therapy for people with cystic fibrosis. Specific research questions were: What are the clinical indications, contraindications and patient selection criteria for non-invasive ventilation use as an adjunct to physical therapy in people with cystic fibrosis? 2. Who implements non-invasive ventilation, what settings are used and how are they determined? 3. What outcome measures are used to determine the effectiveness of non-invasive ventilation as an adjunct to physical therapy and what are the main benefits and complications?

**METHODS:** A purpose-designed survey was sent to 23 Australian cystic fibrosis centres.

**RESULTS:** Fifteen centres (65%) responded, with 13 reporting current utilization of non-invasive ventilation to assist physical therapy. Non-invasive ventilation was most commonly (85%) used in patients with lung function <40% predicted. Physical therapy clinical indications included shortness of breath at rest (100%) and during airway clearance (100%), and fatigue during airway clearance (100%). Physical therapists were responsible for initiating non-invasive ventilation (62%), setting up (85%) and determining settings (62%). Bi-level ventilation was the only chosen ventilation mode. Benefits reported included improved ease of airway clearance (100%), reduced fatigue (92%) and decreased dyspnoea (85%). Only one complication of haemoptysis was reported.

**CONCLUSIONS:** Non-invasive ventilation was used during physical therapy in people with cystic fibrosis who had severe disease, mostly during airway clearance to improve tolerability of treatment. Australian physical therapists initiated non-invasive ventilation when people with cystic fibrosis experienced shortness of breath or fatigue during treatment, aligning with current clinical guidelines. Clinical usage was largely consistent across centres, with numerous benefits and few complications reported. Further research is required to explore benefits of non-invasive ventilation use during physical therapy.

## Introduction

Cystic fibrosis (CF) is a common life-shortening, genetic disease with a prevalence of one in every 2,500 births in Australia (https://www.cysticfibrosis.org.au/about-cf/what-is-cf#What, Accessed March 12, 2019).^1^ Caused by malfunction of a transmembrane conductance regulator protein, CF leads to the development of excessive amounts of thick secretions in the lungs, liver and pancreas (https://www.cff.org/What-is-CF/About-Cystic-Fibrosis/, Accessed March 12, 2019).^2^ People with CF experience recurrent infections, chronic cough, dyspnea and respiratory muscle fatigue as a result of viscous sputum retention and chest wall hyperinflation (https://www.cff.org/What-is-CF/About-Cystic-Fibrosis/, Accessed March 12, 2019).^1,3^ These physiological changes often lead to respiratory failure^4,5^, which is the primary cause of mortality in the CF population.^1^

Despite the severity of the disease, developments in the management of people with CF have improved survival rates^2^; increasing to a median age of 36 years in 2017, from 28 years in 2014.^6,7^ In a 2017 systematic review, Moran et al. suggested that non-invasive ventilation (NIV), a form of positive pressure delivered through a mask^8^, may be an effective tool for slowing the progression of respiratory failure in people with CF. Use of NIV in the CF population, was first described in the literature as a bridge to lung transplantation, with the aim of providing extra time to locate donor lungs.^9^ Since then, the role of NIV has expanded.^8^ NIV is now utilized by physical therapists as an adjunct to airway clearance and exercise^1,10^ and has been proven to decrease fatigue during airway clearance^11^ and reduce oxygen desaturation during exercise.^12^

Many other benefits for the use of NIV in people with CF have been discussed in the literature, whilst few complications have been reported. Suggested advantages for NIV use during physical therapy include reduced shortness of breath (SOB), prevention of small airway collapse, increased tidal volume and reported patient preference.^1,11,13,14^ Reported complications of NIV include mask discomfort, pneumothoraces, air swallowing and drying of secretions.^14,15^ Appropriately fitted masks, use of low pressures and humidification help to minimise the risk of these complications.^1,14,16^ Although the benefits of NIV appear to outweigh the risks, very little literature has been published on how NIV is used in physical therapy for people with CF.

Details regarding the clinical use of NIV to assist airway clearance and exercise for people with CF are not well documented. In fact, a recent study highlighted the lack of validated NIV initiation criteria for people with CF.^17^ It is also unclear whether the decision to implement NIV as an adjunct to treatment is made by the physical therapist, or by other members of the multidisciplinary team. Three previous survey studies have explored NIV use in the CF population. However, none have specifically investigated the use of NIV as an adjunct to airway clearance or exercise. Two studies explored the use of NIV to manage respiratory failure.^18,19^ The third investigated the role of physical therapists and the rationale for NIV use.^20^ Due to the lack of evidence to guide clinical decision making and practicality of use in this area, NIV may be underutilized despite the many proposed advantages.^21^ Therefore, the aim of this study was to investigate the use of NIV by Australian physical therapists, during airway clearance and exercise for people with CF.

The research questions were:

1. What are the clinical indications, contraindications and patient selection criteria for NIV use as an adjunct to physical therapy in people with CF?
2. Who implements NIV, what settings are used and how are they determined?
3. What outcome measures are used to determine the effectiveness of NIV as an adjunct to physical therapy and what are the main benefits and complications?

## Methods

### Design

An electronic survey was developed on Survey Monkey by senior physical therapy staff at a tertiary hospital in Brisbane. The survey, consisting of 33 closed questions and 2 open questions, was distributed to CF centres across Australia. To minimise the time commitment required from participants, the survey was purposefully kept short (i.e. 20-30 minutes). Ethics approval was obtained from the Prince Charles Hospital Human Research Ethics Committee, Metro North Hospital and Health Service obtained prior to the commencement of the study (HREC/16/QPCH/254).

### Procedures

Australian CF centres were eligible for inclusion in the study if identified as an established CF centre in the Australian Cystic Fibrosis Data Registry Annual Report 2016. After CF centres were identified, physical therapy department contact information for each centre was obtained online. Adult, pediatric and mixed centres were included.

Questions for the survey were created after a review of the literature to identify relevant focus areas. Two open ended questions were included to allow centres the opportunity to provide additional information about NIV parameter settings and use in their centre. The survey was pilot tested on four members of the multidisciplinary team (physical therapists, medical and nursing staff) from the investigating facility. One senior physical therapist at the investigating facility did not participate in the pilot testing to ensure availability to complete the survey on behalf of the facility. Based on the feedback provided, the final survey was designed and distributed.

The survey was sent to each CF centre in Australia in November 2016 and requested to be completed by a senior or dedicated CF physical therapist currently working in the centre. One physical therapist from each centre, including the investigating facility, received an individual link to the survey via email, with a unique identification number. The unique identification number was used by the researchers to track which centres had completed the survey. The unique identification number was not used for any other purpose and was not connected to the data. The study involved no foreseeable harm to participants, and therefore consent from the physical therapists was implied once they submitted the survey. This was explained in the survey introduction.

### Participants, therapists, centers

A total of 23 established CF centres were identified and invited to take part in the study. The participating physical therapist was asked to complete the survey on behalf the whole centre to ensure that an overview of current practice in the centre was obtained, rather than the opinions of the individual physical therapist. The planned sample size was consistent with previous similar studies regarding NIV use, where participant numbers ranged from 15 to 36 centres.^18-20^ Participants were given twelve weeks to finalise survey completion, which was deemed sufficient time to retrieve relevant information. To optimise response rate, fortnightly reminder emails were sent to all non-responding centres for the duration of the twelve-week period.

### Data analysis

Data collected on Survey Monkey were stored at the investigating facility in a password protected excel file on a password protected server. After the survey had closed, data were de-identified, assigned a participant number and then exported into Microsoft excel version 1909 (https://support.microsoft.com/en-us/office/what-s-new-in-excel-for-microsoft-365-5fdb9208-ff33-45b6-9e08-1f5cdb3a6c73, October 4, 2019). Descriptive analysis of quantitative data, including frequencies and percentages, were completed in Microsoft excel. Qualitative data from the two open ended questions were analysed manually for content clarification and included in the relevant sections.

### Results

### Flow of participants through the study

Fifteen centres submitted a completed survey, representing a response rate of 65%. Demographic information for the adult (n = 7), pediatric (n = 6) and mixed (n = 2) centres, including number of CF patients and number of inpatients, is presented in Table 1. Thirteen of the 15 (87%) centres reported current utilization of NIV as a treatment option to facilitate airway clearance and/or exercise for people with CF. The two centres that did not currently utilize NIV were pediatric centres, with between 100 and 200 patients. Both reported the lack of use was due to a lack of appropriate patients. These two pediatric centres were excluded from further analysis as the centres were not able to comment on NIV use.

**Table 1.** Characteristics of responding cystic fibrosis centers and the reported number of patients who have utilized non-invasive ventilation as an adjunct to physical therapy in the past 12 months presented as number (%)

|  | Adult Centers | Pediatric Centers | Mixed Centers |
| --- | --- | --- | --- |
| **No. of centers responding to survey (n = 15)** | 7 (47) | 6 (40) | 2 (13) |
| **Total no. of CF patients managed by responding center** |  |  |  |
| Between 20-150 | 2 (13) | 2 (13) | 0 (0) |
| Between 150-250 | 3 (20) | 3 (20) | 2 (13) |
| More than 250 | 2 (13) | 1 (7) | 0 (0) |
| **No. of centers using NIV** | 7 (47) | 4 (27) | 2 (13) |
| Reported frequency of NIV use in the past 12 months (n = 13) |  |  |  |
| **How many inpatients have utilized NIV in the past 12 months at your center?** |  |  |  |
| With airway clearance |  |  |  |
| None | 0 (0) | 1 (8) | 0 (0) |
| Between 1-6 | 3 (23) | 1 (1) | 1 (8) |
| Between 7-15 | 0 (0) | 2 (15) | 0 (0) |
| More than 16 | 4 (31) | 0 (0) | 1 (8) |
| With exercise |  |  |  |
| None | 3 (23) | 1 (8) | 0 (0) |
| Between 1-6 | 4 (31) | 3 (23) | 1 (8) |
| Between 7-15 | 0 (0) | 0 (0) | 1 (8) |
| More than 16 | 0 (0) | 0 (0) | 0 (0) |
| **How many outpatients** |  |  |  |
| **have utilized NIV in the** |  |  |  |
| **past 12 months at your** |  |  |  |
| **center?** |  |  |  |
| With airway clearance |  |  |  |
| None | 2 (15) | 2 (15) | 0 (0) |
| Between 1-6 | 5 (38) | 2 (15) | 1 (8) |
| Between 7-15 | 0 (0) | 0 (0) | 0 (0) |
| More than 16 | 0 (0) | 0 (0) | 1 (8) |
| With exercise |  |  |  |
| None | 4 (31) | 4 (31) | 0 (0) |
| Between 1-6 | 3 (23) | 0 (0) | 1 (8) |
| Between 7-15 | 0 (0) | 0 (0) | 1 (8) |
| More than 16 | 0 (0) | 0 (0) | 0 (0) |

CF = cystic fibrosis
NIV = non-invasive ventilation

### Usage of NIV

Of the 13 CF centres that implemented NIV as an adjunct to physical therapy, 11 (85%) reported that NIV was most commonly used in people who had a forced expiratory volume in one second (FEV_1_) of <40% predicted. Two centres (15%) reported that NIV was most commonly used in people with CF who had an FEV_1_ of 40-70% predicted. Six of the 13 centres that utilized NIV (46%) reported there was a formal training requirement for physical therapists using NIV during airway clearance or exercise in people with CF in their centre. In addition, five centres (38%) indicated the presence of a current policy or guideline for the use of NIV as an adjunct to physical therapy in the CF population. Table 1 presents the reported number of inpatients and outpatients that had utilized NIV as an adjunct to physical therapy in the past 12 months. Table 2 presents the most frequently reported clinical indications, contraindications and patient selection criteria for the use of NIV as an adjunct to physical therapy in people with CF.

**Table 2.** Clinical indications, contraindications and patient selection criteria for non-invasive ventilation use as an adjunct to physical therapy presented as number (%)

|  | N (%) |
| --- | --- |
| **Indications** |  |
| Shortness of breath at rest | 13 (100) |
| Shortness of breath during airway clearance | 13 (100) |
| Fatigue during airway clearance | 13 (100) |
| Difficulty clearing secretions | 12 (92) |
| **Contraindications** |  |
| Recent/undrained pneumothorax | 8 (62) |
| Active haemoptysis | 7 (54) |
| Upper airway/oesophageal/gastric surgery | 2 (15) |
| **Patient selection criteria** |  |
| Airway clearance |  |
| Fatigue during usual airway clearance | 12 (92) |
| Difficulty clearing secretions | 9 (69) |
| Hypercapnia | 5 (38) |
| Patient comfort | 5 (38) |
| Exercise |  |
| Shortness of breath at rest | 10 (77) |
| Shortness of breath with minimal exertion | 9 (69) |
| Fatigue during minimal exertion | 9 (69) |
| Hypoxia at rest | 4 (31) |

NIV = non-invasive ventilation

### Indications

Figure 1 presents reported clinical indications for the use of NIV as an adjunct to physical therapy in people with CF. Six respondents (46%) reported other indications, including post spontaneous rib fractures in patients who had difficulty with inspiration (n = 1). Indication for NIV use is based on individual assessment of each patient, with one centre reporting that clinical indicators would not automatically trigger NIV use (n = 1). Acute hypercapnic respiratory failure, hypoxaemia and use as a bridge to transplantation were also identified (n = 1) as important indications for long-term implementation of NIV, which is managed by medical and nursing staff.

**Figure 1.**
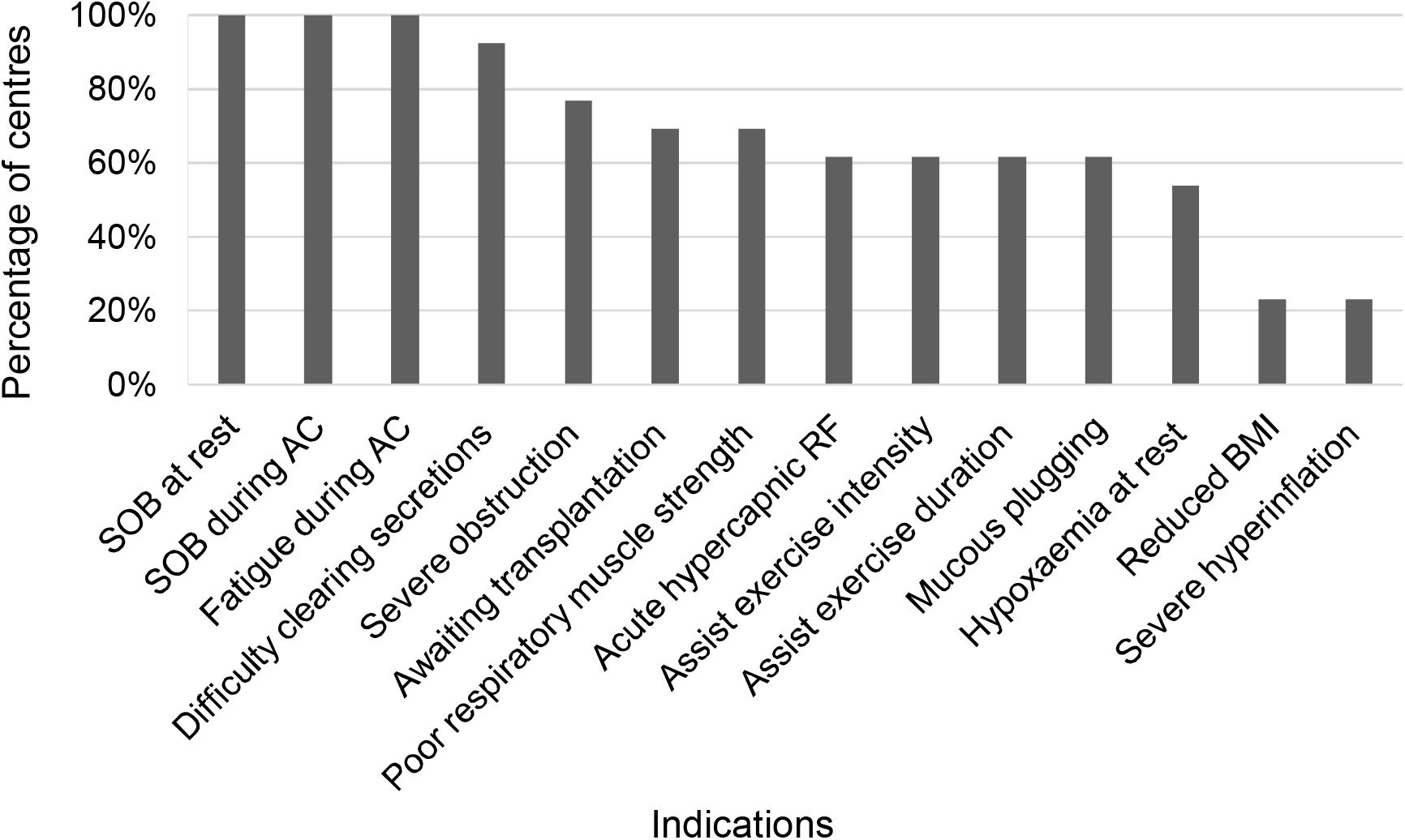
Clinical indications for the use of non-invasive ventilation in people with cystic fibrosis

### Contraindications

Centres were asked to identify contraindications in addition to those listed in the British Thoracic Society Guidelines.^22^ Ten (77%) centres reported specific contraindications to the use of NIV during physical therapy for people with CF in their centre. The most common contraindication was a recent or undrained pneumothorax, reported by eight centres (62%). Seven centres (54%) identified that an active, new or significant haemoptysis would be considered a contraindication for implementing NIV in a person with CF. Other contraindications identified by responding centres included aerophagia (n = 1), upper airway surgery (n = 1), inability to protect airway (n = 1), life-threatening hypoxaemia (n = 1), base of skull fractures (n = 1), rib fractures (n = 1), uncontrolled vomiting (n = 2) and recent oesophageal or upper gastrointestinal surgery (n = 2) (Table 2).

### Patient selection criteria

The most important patient selection criterion for NIV use to assist airway clearance, was fatigue during usual airway clearance, identified by 92% (n =12) of centres. Other commonly reported patient selection criteria for NIV use to assist airway clearance included difficulty clearing secretions (69%), patient comfort (38%) and hypercapnia (38%). The most important patient selection criteria for NIV use during exercise were shortness of breath (SOB) at rest (77%) and SOB with minimal exertion (69%). Nine centres (69%) identified that fatigue during minimal exertion was used to select people with CF for NIV use during exercise. In addition, hypoxia at rest was chosen as an important criterion in 31% of centres. One centre commented that CF patients on veno-venous extracorporeal membrane oxygenation who are bridging to lung transplant may be selected for NIV use during exercise.

### Implementation of NIV

Eight centres (62%) reported that a physical therapist was always responsible for initiating the use of NIV as an adjunct to physical therapy. Conversely, 69% of centres reported that the CF medical specialist was occasionally responsible for initiating NIV as an adjunct to physical therapy. Ten centres (77%) identified that the respiratory nurse was never responsible for initiating NIV as an adjunct to physical therapy. In 11 (85%) of the responding centres, a physical therapist was responsible for the set up and application of an initial trial of NIV to assist physical therapy treatment in a person with CF. Eight centres (62%) identified that NIV settings were usually determined by a physical therapist, whilst 31% (n = 4) stated that settings were determined by a medical team member. One centre specified that decisions about NIV settings were made in consultation with the whole team (Table 3).

**Table 3.** Implementation of non-invasive ventilation, settings and factors used to determine the settings presented as number (%)

|  | N (%) |
| --- | --- |
| Who is responsible for the set up and application of NIV? |  |
| Physical therapist | 11 (85) |
| Other | 2 (15) |
| Who are the setting parameters determined by? |  |
| Physical therapist | 8 (62) |
| Other | 5 (38) |
| **Settings** |  |
| Ventilation mode |  |
| Bi-level Positive Airway Pressure | 13 (100) |
| IPAP settings |  |
| <10cmH<sub>2</sub>O | 1 (8) |
| 10-15cmH<sub>2</sub>O | 10 (77) |
| 15-20cmH<sub>2</sub>O | 2 (15) |
| EPAP settings |  |
| <10cmH<sub>2</sub>O | 13 (100) |
| Interfaces |  |
| Full face mask | 6 (46) |
| Mouthpiece | 4 (31) |
| Nasal mask | 2 (15) |
| Humidification |  |
| Always | 9 (70) |
| Based on individual assessment | 2 (15) |
| **Reasons for increasing IPAP \*** |  |
| Lack of subjective benefit by patient | 10 (77) |
| Lack of perceived benefit by therapist | 9 (69) |
| Severity of dyspnoea at rest | 8 (61) |
| Patient tolerance of pressure | 8 (61) |
| Larger patient size | 7 (54) |
| **Reasons for decreasing IPAP \*** |  |
| Patient discomfort | 11 (85) |
| Therapist concern for complication | 10 (77) |
| Less severe pulmonary obstruction | 2 (15) |
| **Determining EPAP \*** |  |
| Requirement for airway splinting | 12 (92) |
| Ability to perform forced expiration | 8 (61) |
| Patient comfort | 8 (61) |
| Presence of mucous plugging | 7 (54) |
| Therapist concern for complication | 7 (54) |
| **Determining interface \*** |  |
| Patient comfort | 11 (85) |
| Requirement for splinting of airways | 4 (31) |
| Patient need for expectoration | 3 (23) |
| Mask availability | 2 (15) |

NIV = non-invasive ventilation
IPAP = inspiratory positive airway pressure
EPAP = expiratory positive airway pressure
\*multiple answers

### Ventilation and parameter settings

Ventilation and parameter settings including the most common ventilation mode, pressures, and interfaces used by physical therapists are presented in Table 3. One centre (8%) reported setting expiratory pressures in a similar way to positive expiratory pressure devices, increasing pressures to 10-15cmH_2_O for a few sets with resting sets in between. Another centre commented that they commonly increased expiratory pressures during airway clearance treatment, for example, from 7cmH_2_O pre-treatment to 11cmH_2_O during treatment.

Patient comfort was considered an important factor when determining NIV settings in 85% of centres (n = 11). Other commonly reported factors included pulse oximetry (69%) and observation of patient-ventilator synchrony (62%). Centres also identified that patient feedback (n = 1), clinical observation (n = 1) and current nocturnal settings (n = 1) were often used to determine NIV settings for physical therapy. One centre reported that initial settings were determined by a consultant and titrated by a physical therapist as required. Reasons for increasing or decreasing inspiratory positive airway pressure, and factors used to choose the expiratory positive airway pressure and interface are presented in Table 4.

**Table 4.** Outcome measures used to determine effectiveness of non-invasive ventilation use and associated benefits and complications presented as number (%)

|  | N (%) |
| --- | --- |
| **Outcome measures used** |  |
| In a single airway clearance session |  |
| Decreased fatigue | 13 (100) |
| Patient subjective benefit | 12 (92) |
| Increased ease of sputum clearance | 12 (92) |
| Increased tolerability of treatment | 12 (92) |
| Decreased shortness of breath | 12 (92) |
| In a single exercise session |  |
| Patient subjective benefit | 13 (100) |
| Improved exercise endurance | 13 (100) |
| Increased exercise intensity | 12 (92) |
| Decreased work of breathing | 11 (85) |
| Over the course of a patient's admission |  |
| Patient subjective benefit | 13 (100) |
| Observation of breathing | 12 (92) |
| Oxygen saturations | 10 (77) |
| Respiratory rate | 10 (77) |
| **Benefits of NIV use** |  |
| Improved tolerance of airway clearance | 13 (100) |
| Ease of sputum clearance | 13 (100) |
| Reduced fatigue | 12 (92) |
| Decreased dyspnoea | 11 (85) |
| Improved exercise tolerance | 8 (62) |
| **Reasons for failure of NIV use** |  |
| Absence of benefit as determined by therapist | 9 (69) |
| Patient discomfort | 8 (62) |
| Absence of subjective benefit for patient | 6 (46) |
| Patient or carer wish to withdraw treatment | 4 (31) |
| **Complications with NIV use** |  |
| None | 12 (92) |
| Haemoptysis | 1 (8) |
| Pneumothorax | 0 (0) |
| **Problems with NIV use** |  |
| No subjective benefit | 6 (46) |
| None | 4 (31) |
| Problems with interface | 4 (31) |

NIV = non-invasive ventilation

### Outcome measures, benefits, problems and complications

Table 4 presents outcome measures used to determine NIV effectiveness, associated benefits and complications. Level of fatigue was reportedly used as an outcome measure to assess effectiveness of NIV use during a single airway clearance session in 100% (n = 13) of centres. Other commonly reported outcome measures included patient reported benefit, ease of clearance, tolerability of treatment and SOB, identified by 92% (n = 12) of centres. Sputum volume (77%) and oxygenation (77%) were also used in the responding centres. During a single session of exercise, outcome measures reported to determine effectiveness of NIV included patient reported benefit (100%), exercise endurance (100%) and intensity (92%), level of fatigue (77%), oxygenation (77%) and SOB (69%).

Improved tolerance/ease of airway clearance was the most common benefit, reportedly observed in 100% of responding centres. Other frequently reported benefits were reduced fatigue (92%), decreased dyspnoea (85%) and improved exercise tolerance (62%). Improved partial pressure of oxygen (46%) and reduced partial pressure of carbon dioxide (38%) were also reported following NIV use in some centres. One centre reported improvements in sleep, relaxation, heart rate and mucolytic effect were observed in their centre.

NIV use was reportedly discontinued in the absence of a benefit perceived by the therapist (69%), claustrophobia (62%), and failure to alleviate symptoms (46%). Problems associated with NIV use included a lack of subjective benefit reported by the patient, identified in 46% (n = 6) of centres, and problems with the interface, identified in 31% (n = 4) of centres. The only reported complication that occurred with NIV use was haemoptysis (n = 1), and the centre proposed that the haemoptysis was not necessarily caused by NIV. In addition, 92% (n = 12) of the responding centres reported that none of their patients had experienced a complication whilst using NIV during physical therapy.

## Discussion

NIV was implemented as an adjunct to physical therapy in adult, pediatric and mixed CF centres across Australia. However, NIV was only used with low numbers of patients, the majority of whom had severe lung disease. NIV was also more frequently used to assist airway clearance than exercise. Physical therapists implemented NIV with patients who experienced fatigue during airway clearance treatment. During exercise, NIV was also used in those who experienced fatigue, as well as those experiencing SOB, either at rest or with minimal exertion. In Australia, physical therapists were frequently involved in the set up and application of NIV for physical therapy treatment, as well as determining settings. Ventilator settings used appear to be similar across Australian centres, with multiple factors, such as patient comfort and severity of dyspnoea, determining the settings. Numerous benefits and few complications associated with NIV use during physical therapy were reported.

Responding centres indicated that NIV was most frequently implemented in people with severe CF lung disease (i.e. FEV_1_ <40% predicted) and that NIV may increase tolerability of physical therapy treatment. Previous studies have also identified that NIV was most commonly used in those with severe lung disease (FEV_1_ of 22-35% predicted).^18^ Increased dyspnoea and respiratory muscle fatigue caused by severe lung disease may limit effective airway clearance and exercise in people with CF.^10^ Therefore, people with more severe disease are likely to benefit from respiratory support, such as NIV, during physical therapy sessions.^10^ In fact, research indicates that NIV can improve secretion clearance^13^, and decrease dyspnoea^14^ and fatigue^11^ during airway clearance treatment in those with moderate to severe CF. This may explain why NIV use was less common in pediatric centres, as lung disease in children and adolescents is usually less severe than during adulthood due to the progressive nature of the disease.^23^

Clinical guidelines from the United Kingdom suggest that NIV may be used to assist airway clearance in people with CF who have difficulty clearing secretions.^24^ The Australian and New Zealand guideline states NIV is a useful adjunct to airway clearance in people with CF who experience dyspnoea and fatigue.^10^ Findings from the current study indicate that Australian physical therapists were using NIV in line with these recommendations. The main clinical indications were SOB at rest, and SOB and fatigue during airway clearance. Difficulty clearing secretions was also reportedly used to select patients for NIV use during airway clearance. Our findings align with a previous survey where 68% of respondents reported NIV was a valid tool to support airway clearance techniques in people with CF.^20^

NIV was more commonly used to assist airway clearance compared to exercise in Australian centres. This may be due to airway clearance being the cornerstone of physical therapy management for people with CF due to recurrent infection.^10,24^ Respondents were working in acute inpatient hospital settings, where NIV may be utilized to manage symptoms of an acute exacerbation.^11^ This may account for the more frequent reported use of NIV to assist airway clearance for inpatients. In addition, more evidence exists for airway clearance and may contribute to the increased use in this area.^11,13,14,25^

Despite reported benefits for NIV use during exercise^12^, NIV was rarely used by responding centres for this purpose. Clinical guidelines from Australia and the United Kingdom support the use of NIV during exercise to reduce SOB, increase oxygenation and improve exercise tolerance.^10,24^ However, the evidence supporting these guidelines is limited.^12^ When selecting patients to use NIV during exercise, responding centres reported SOB at rest or during minimal exertion, fatigue during minimal exertion and hypoxia, as the most important criteria. More than half of the centres in the current study also reported that improved exercise tolerance was observed in their patients after NIV use. Although it appears that NIV use during exercise is indicated for people with CF and may be beneficial, further research is required to inform clinical practice.

More than half of the responding centres did not report a formal training requirement or current policy for NIV use in physical therapy. Regardless, NIV settings used appear to be consistent between CF centres in Australia. Australian physical therapists most commonly set inspiratory pressure between 10-15cmH_2_O, with a maximum of 20cmH_2_O. Similar inspiratory pressures were used in two Australian randomised trials, ranging from 10 to 20cmH_2_O.^11,14^ This contrasts with European findings, where maximum inspiratory pressures of 32cmH_2_O have been used.^13,25^ This suggests that inspiratory pressure settings used in Australia may be more conservative than those used in Europe. Expiratory pressures reported in the current study were more consistent with those used in the literature. All respondents in the current study used expiratory pressures of less than 10cmH_2_O. This aligns with British and Australian studies where expiratory pressures of less than 8cmH_2_O were used.^11,13,14^ Interestingly, higher expiratory pressures ranging from 12 to 14cmH_2_O, have been used in a recent study with no associated complications.^25^

Patient comfort was frequently used by physical therapists to determine NIV settings. This suggests that subjective feedback plays an important role in the clinical reasoning process, with the aim of improving acceptance of treatment. In adult and pediatric CF centres in France, 73-92% of respondents also reported patient comfort determined NIV settings for people with CF in respiratory failure.^18^ Our study suggests that inspiratory pressures were set based on patient perceived benefit, and severity of dyspnoea at rest. These factors could be utilized by clinicians to further support clinical decision making regarding the amount of respiratory support required to decrease SOB and unload the respiratory muscles.^26^ It appears that expiratory pressures were selected based on the minimum pressure required for splinting collapsible airways, whilst ensuring the patient was comfortable and able to perform forced expiration. Overall, centres appeared to use the lowest pressures possible to splint the airways, decrease SOB and increase gas exchange.^1,26,27^

Importantly, few complications were associated with the use of NIV during physical therapy in the current study. Complications, such as air swallowing and pneuomothoraces, have been reported in the past.^15,19^ Skin breakdown^27^ and secretion retention have also been identified.^19^ However, these complications are often related to long-term NIV use for management of respiratory failure and are rarely seen in the application of NIV during physical therapy.^16^ It is possible that conservative application of NIV and the use of relatively low pressure settings are contributing factors to less incidence of complications.

During the current study, strategies such as fortnightly reminder emails were put in place to optimise response rate. Despite these, approximately one-third of Australian centres did not respond. Additionally, two respondents did not currently utilize NIV as an adjunct to physical therapy for people with CF. Although this sample size was consistent with previous studies regarding NIV use^18-20^, it was disappointing that more centres did not respond. Regardless, respondents included adult, pediatric and mixed CF centres of various sizes and were representative of most states and territories in Australia. A further limitation of this study was that the survey was only distributed to CF centres in Australia. Therefore, few comparisons between countries regarding the clinical indications and implementation of NIV during physical therapy could be made. Although centres were asked to answer questions on behalf of the centre, personal opinions may have had an impact on some results.

### Conclusion

This is the first study to investigate the use of NIV by Australian physical therapists for people with CF and explore whether clinical use is in line with the evidence. Findings revealed that centres were using NIV as an adjunct to physical therapy in low numbers of people with CF. NIV was more commonly used in adult centres, in those with severe CF lung disease and to assist airway clearance. Reported indications for the use of NIV during physical therapy appear to align with Australian and British clinical guidelines for physical therapy management of CF. Physical therapists were frequently responsible for initiating NIV and determining NIV settings, which were consistent across CF centres in Australia. Australian centres appear to be conservative when setting inspiratory pressures and reported that complications associated with NIV use were rare. More rigorous randomised controlled trials should be conducted in the future to provide further evidence for the benefits of NIV use during airway clearance and exercise for people with CF.

## Data Availability

The datasets generated during and/or analysed during the current study are not publicly available, however are available from the corresponding author on reasonable request.

## Acknowledgements

We would like to thank the responding centres for their contribution to this research.

## Appendices

### Appendix 1: Survey

The Use of Non-Invasive Ventilation as an Adjunct to Physical therapy in Cystic Fibrosis Patients – A Survey of Physical therapists

It is anticipated that this survey will take 20-30 minutes to complete, so please ensure you have this amount of time to fully complete it.

Please complete this survey even if you have no or limited experience with non-invasive ventilation (NIV).

Please ensure only one survey is completed by your centre.

Your participation in this survey is voluntary. If you agree to participate, please be assured that your responses will remain confidential. Your consent to participate will be implied by completion of the online survey.

Please note that the link you clicked on is unique to your cystic fibrosis (CF) centre. This is to allow us to follow up with centres that have not yet responded to the survey. These unique identifications will only be used for this purpose, and will not be linked to answers or be used for any other purpose during the study.

For the purposes of this survey, “NIV as an adjunct to physical therapy” encompasses the use of NIV to facilitate or assist airway clearance and/or exercise. This survey does not relate to patient use of NIV for continuous ventilatory support, nocturnal ventilation or management of respiratory failure.

All questions relate to the use of NIV as an adjunct to physical therapy in patients with cystic fibrosis only – not the use of NIV in any other lung conditions

### Background Information

1. Does your facility provide Adult or Pediatric CF services?
  ◯ Adult
  ◯ Pediatric
  ◯ Combination of Adult and Pediatric services
2. How many CF patients does your facility care for?
  ◯ Less than 20 patients
  ◯ 20 - 50 patients
  ◯ 100 - 150 patients
  ◯ 150 - 200 patients
  ◯ 200 - 250 patients.
  ◯ More than 250 patients
3. On average, how many CF patients do you have in hospital at any one time?
  ◯ Rarely have any inpatients\
  ◯ 1 – 3 inpatients
  ◯ 4 – 6 inpatients
  ◯ 7 – 10 inpatients
  ◯ 11 – 15 inpatients
  ◯ 16 – 20 inpatients
  ◯ More than 20 inpatients
4. Does your facility have a formal training requirement or competency for physical therapists who utilize NIV as an adjunct to physical therapy for CF patients?
  ◯ Yes
  ◯ No
5. Does your facility have a current policy or guideline for NIV as an adjunct to physical therapy for CF patients?
  ◯ Yes
  ◯ No
6. Does your facility currently utilize non-invasive ventilation (NIV) as a treatment option to facilitate airway clearance and/or exercise for CF patients?
  ◯ Yes
  ◯ No
7. Please identify the reasons your facility does not utilize NIV as a treatment option to facilitate airway clearance and/or exercise for CF patients. Please select all that apply.
  ◯ Not required
  ◯ Lack of evidence to support its use
  ◯ Lack of staff expertise
  ◯ Lack of equipment
  ◯ Lack of staff resourcing
  ◯ Lack of appropriate patients
  ◯ Lack of support from medical team
  ◯ Other, please specify

#### Indications for NIV Use

8. For what indication/s do you utilize NIV as an adjunct to physical therapy in CF patients? Please select all that apply
  ◯ Acute hypercapnic respiratory failure
  ◯ Hypoxaemia at rest
  ◯ Severe impairment of pulmonary function/ Severe airflow obstruction
  ◯ Difficulty clearing secretions/poor forced expirations
  ◯ Shortness of breath/increased work of breathing at rest Shortness of breath during airway clearance
  ◯ Fatigue during airway clearance
  ◯ Assist exercise training intensity
  ◯ Assist exercise training duration
  ◯ Awaiting lung transplantation
  ◯ Poor respiratory muscle strength
  ◯ Reduced BMI/body weight\
  ◯ Severe pulmonary hyperinflation
  ◯ Mucous plugging identified by chest x-ray or CT scan
  ◯ Other, please specify
9. Based on your clinical experience, what are the <u>most important</u> criteria for patient selection for successful and effective NIV use <u>to assist airway clearance</u> in CF patients? **Please select <u>maximum of 3 options</u>**
  ◯ Fatigue during usual airway clearance technique
  ◯ Hypoxaemia
  ◯ Hypercapnia
  ◯ Patient willingness to trial/use
  ◯ Mucous plugging identified by chest x-ray or CT scan
  ◯ Patient comfort
  ◯ Patient subjective opinion of benefit
  ◯ Difficulty clearing secretions
  ◯ Other, please specify
10. Based on your clinical experience, what are the <u>most important</u> criteria for patient selection for successful and effective NIV use <u>to assist exercise</u> in CF patients? **Please select <u>maximum of 3 options</u>**
  ◯ Shortness of breath at rest
  ◯ Shortness of breath with minimal exertion
  ◯ Shortness of breath with usual exercise
  ◯ Fatigue during usual exercise
  ◯ Fatigue during minimal exertion
  ◯ Hypoxia at rest
  ◯ Exercise-induced hypoxia
  ◯ Other, please specify
11. Recent BTS/ICS Guidelines (Davidson et al, 2016, Thorax) list the following contraindications to the use of NIV. Absolute contraindications:
  – Severe facial deformity
  – Facial burns
  – Fixed upper airway obstruction Relative contraindications:
  – pH < 7.15
  – GCS < 8
  – Confusion / agitation
  – Cognitive impairment
  Aside from those listed above, at your facility are there any other contraindications to the use of NIV as an adjunct to physical therapy specifically for CF patients?
  ◯ No
  ◯ Yes - please specify

#### Usage

12. At your facility, approximately how many CF **<u>inpatients</u>**<u>in the past 12 months</u> have utilized NIV as an adjunct to physical therapy:

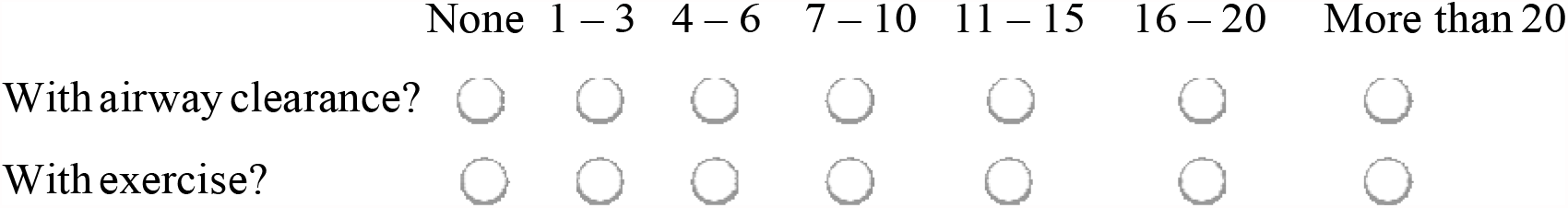
13. At your facility, approximately how many CF **<u>inpatients</u>** have started using NIV as an adjunct to physical therapy for the **<u>first time</u>**<u>in the</u> **<u>past 12 months</u>?**

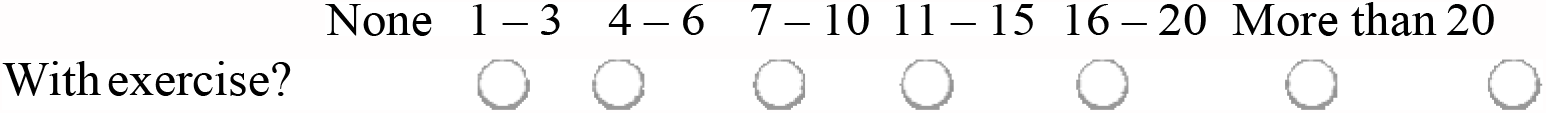
14. At your facility, approximately how many CF **<u>outpatients</u>**<u>in the</u> **<u>past 12 months</u>** have utilized NIV as an adjunct to physical therapy:

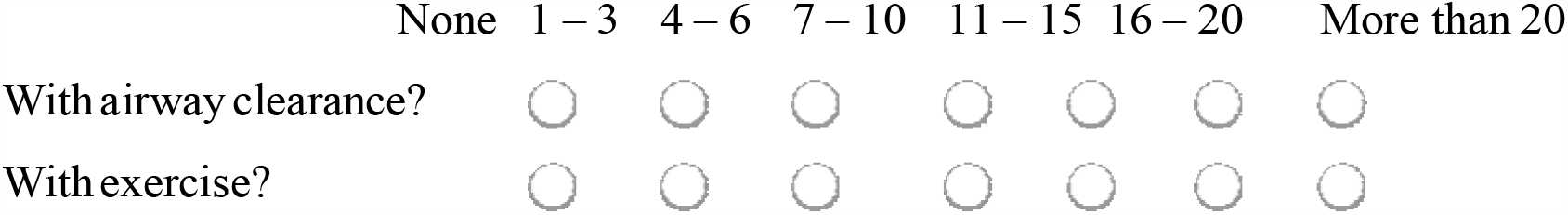
15. Most commonly, patients at your facility who use/have used NIV as an adjunct to physical therapy have an FEV_1_%pred that falls in the following ranges:
  ◯ FEV1%pred greater than 70%
  ◯ FEV1%pred 40% - 70%
  ◯ FEV1%pred less than 40%
  ◯ Cannot accurately answer

#### Team Members

16. At your facility, which CF team member is responsible for suggesting or initiating the use of NIV as an adjunct to physical therapy?

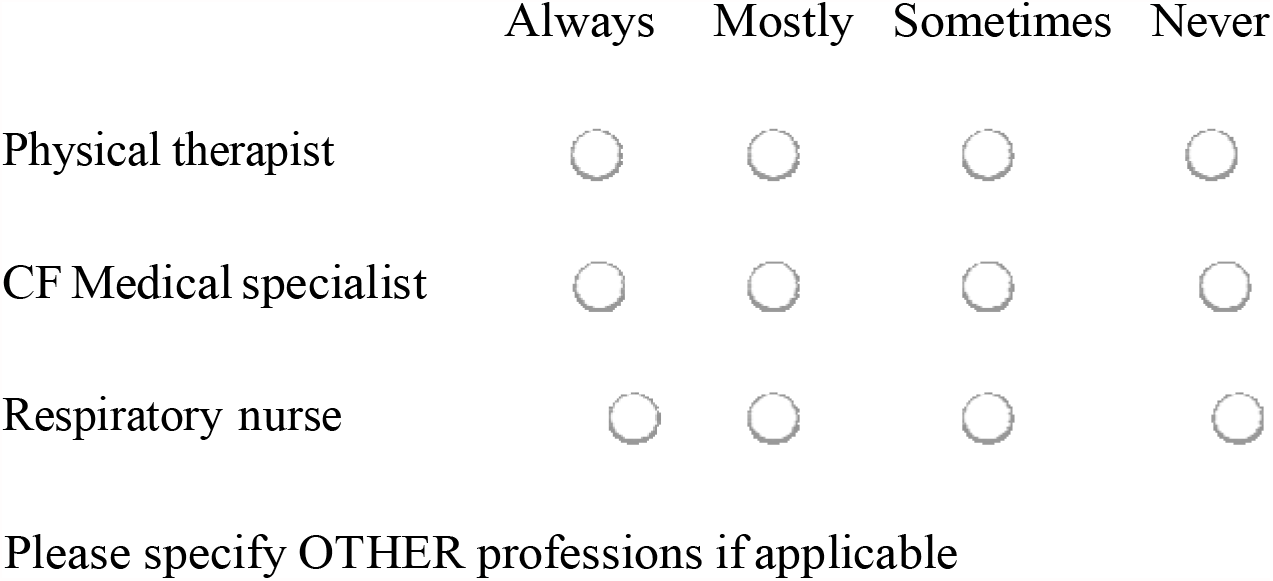
17. For a CF patient’s <u>initial tria</u>l of NIV as an adjunct to physical therapy:

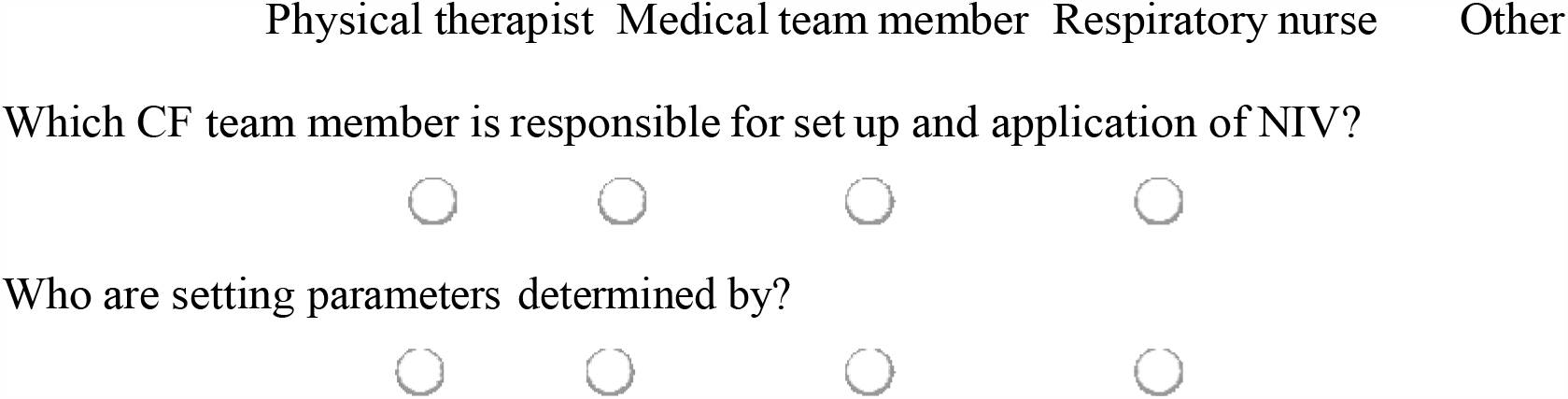 Comments

#### Ventilation and Parameter Settings

18. For the use of NIV as an adjunct to physical therapy in CF patients, which <u>ventilation mode</u> do you most commonly use?
  ◯ Continuous Positive Airway Pressure
  ◯ Bi-level Positive Airway Pressure
  ◯ Volume controlled
  Comments
19. How are the NIV settings determined for a CF patient using NIV as an adjunct to physical therapy?

#### Please select all that apply

◯ Unsure
◯ Patient comfort during NIV
◯ Severity of pulmonary hyperinflation
◯ Severity of pulmonary obstruction
◯ Severity of dyspnoea at rest
◯ Highest tolerated by patient
◯ Pulse oximetry
◯ ABG during NIV
◯ ABG during spontaneous breathing
◯ Patient-ventilator synchrony
◯ Other, please specify

20. Based on your clinical experience, if using bi-level positive airway pressure (BiPAP) as an adjunct to physical therapy, for what reasons would a patient require <u>higher/increasing inspiratory positive airway pressure (IPAP)?</u> **Please select all that apply**
  ◯ Unsure / do not use
  ◯ Patient comfort during NIV
  ◯ Severity of pulmonary hyperinflation
  ◯ Severity of pulmonary obstruction
  ◯ Severity of dyspnoea at rest
  ◯ Patient tolerance of pressure
  ◯ Larger patient size
  ◯ ABG during NIV
  ◯ Gender
  ◯ Medical team request
  ◯ Patient-ventilator synchrony
  ◯ Lack of subjective benefit by patient
  ◯ Lack of perceived benefit by therapist
  ◯ Other, please specify
21. Based on your clinical experience, if using bi-level positive airway pressure (BiPAP) as an adjunct to physical therapy, for what reasons would a patient require <u>lower/decreasing inspiratory positive airway pressure (IPAP)?</u> **Please select all that apply**
  ◯ Unsure / do not use
  ◯ Patient discomfort
  ◯ Less severe pulmonary obstruction
  ◯ Therapist concern for complication
  ◯ Other, please specify
22. If using bi-level positive airway pressure (BiPAP) as an adjunct to physical therapy, how do you determine the level of expiratory positive airway pressure (EPAP) to provide? **Please select all that apply**
  ◯ Unsure / do not use
  ◯ Patient comfort
  ◯ Severity/presence of mucous plugging
  ◯ Requirement for airway splinting/collapsible airways
  ◯ Effect on ability to perform forced expirations/cough
  ◯ Therapist concern for complication
  ◯ Other, please specify
23. If using bi-level positive airway pressure (BiPAP) as an adjunct to physical therapy, what pressure settings are most commonly used at your centre?

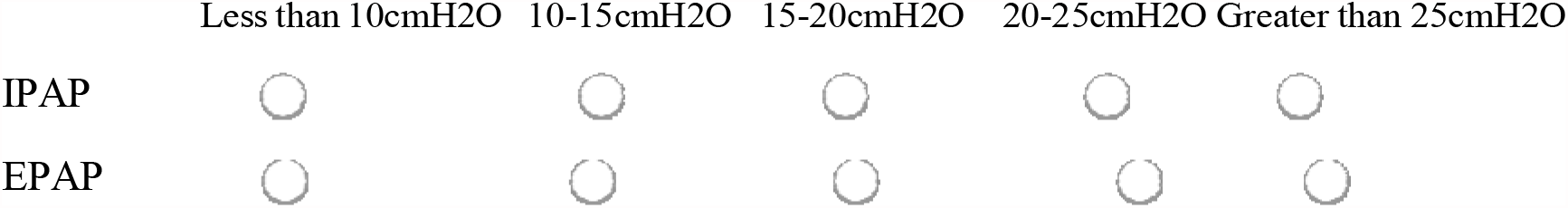
24. Please feel free to give any other comments or description regarding NIV parameter settings

|  | Less than 10cmH<sub>2</sub>O | 10-15cmH<sub>2</sub>O | 15-20cmH<sub>2</sub>O | 20-25cmH<sub>2</sub>O | Greater than 25cmH<sub>2</sub>O |
| --- | --- | --- | --- | --- | --- |
| IPAP | ☐ | ☐ | ☐ | ☐ | ☐ |
| EPAP | ☐ | ☐ | ☐ | ☐ | ☐ |

#### Interface and Humidification

25. For the use of NIV as an adjunct to physical therapy for CF patients, which interface type is most commonly used at your facility?
  ◯ Nasal mask
  ◯ Nasal pillows
  ◯ Full face mask
  ◯ Mouthpiece
  ◯ Other (please specify)
26. For the use of NIV as an adjunct to physical therapy for CF patients at your facility, interface type is usually determined by:
  ◯ Mask availability
  ◯ Patient comfort
  ◯ Patient need for expectoration
  ◯ Requirement for splinting of airways/collapsible airways
  ◯ Other (please specify)
27. For the use of NIV as an adjunct to physical therapy for CF patients at your facility, is humidification used with NIV application?
  ◯ Always
  ◯ Based on individual assessment
  ◯ Never
  ◯ Other (please specify)

#### Outcomes

28. Based on your clinical experience, what benefits do you commonly see with the use of NIV as an adjunct to physical therapy? **Please select all that apply**
  ◯ Improved tolerance of airway clearance
  ◯ Improved exercise tolerance
  ◯ Improvement in PaO2
  ◯ Reduction in PaCO2
  ◯ Less fatigue
  ◯ Decreased dyspnoea
  ◯ Ease of sputum clearance
  ◯ Other (please specify)
29. What outcome measures do you use to determine the effectiveness of the use of NIV to assist a <u>single airway clearance session.</u> **Please select all that apply**
  ◯ Patient subjective benefit
  ◯ Increased volume of sputum
  ◯ Increased ease of sputum clearance
  ◯ Decreased fatigue
  ◯ Increased tolerability of treatment
  ◯ Decreased shortness of breath
  ◯ Improved oxygenation
  ◯ Decreased heart rate
  ◯ Other (please specify)
30. What outcome measures do you use to determine the effectiveness of the use of NIV to assist a <u>single exercise session.</u> **Please select all that apply**
  ◯ Patient subjective benefit
  ◯ Decreased fatigue
  ◯ Decreased shortness of breath/ reduce dyspnoea
  ◯ Improved oxygenation
  ◯ Decreased work of breathing
  ◯ Decreased heart rate response
  ◯ Able to exercise for longer duration/improve exercise endurance
  ◯ Able to exercise at higher intensity
  ◯ Other (please specify)
31. Over the course of the CF patients admission, what measures are routinely used to determine the effectiveness of NIV as an adjunct to physical therapy? **Please select all that apply**
  ◯ Oxygen saturations
  ◯ Arterial blood gases
  ◯ Tidal volume
  ◯ Respiratory rate
  ◯ Heart rate
  ◯ Lung function (FEV1)
  ◯ Observation of patient – breathing pattern, work of breathing
  ◯ Patient subjective perception of benefit
  ◯ Other (please specify)

#### Complications

32. What are the main reasons for a CF patient to fail or prematurely discontinue NIV as an adjunct to airway clearance or exercise? **Please give <u>maximum of 3 options</u>**
  ◯ Absence of a clear subjective benefit for patient/failure to alleviate symptoms
  ◯ Absence of a benefit as determined by therapist.
  ◯ Patient discomfort: claustrophobic feeling of mask
  ◯ Patient discomfort: poor tolerability of heat caused by humidification
  ◯ Patient discomfort: dis-synchrony with ventilator
  ◯ Deterioration of patient condition.
  ◯ Patient or carer wish to withdraw treatment
  ◯ Other (please specify)
33. At your facility, have any CF patients experienced complications that were perceived to be due to the use of NIV as an adjunct to airway clearance or exercise? **Please select all that apply**
  ◯ None
  ◯ Pneumothorax
  ◯ Haemoptysis.
  ◯ Sputum retention
  ◯ Other (please specify)
34. Do CF patients at your facility experience any problems associated with the use of NIV as an adjunct to physical therapy? **Please select all that apply**
  ◯ None
  ◯ Preference for oxygen therapy
  ◯ No subjective benefit
  ◯ Too constraining
  ◯ Problems with interface
  ◯ Refusal
  ◯ Abdominal distension
  ◯ Skin deterioration / pressure areas
  ◯ Other, please specify
35. Please feel free to make any other comments regarding the use of NIV as an adjunct to physical therapy for CF patients.

## QUICK LOOK

**Current knowledge**

Use of non-invasive ventilation (NIV) in the cystic fibrosis (CF) population was first described as a bridge to lung transplantation. Currently, NIV is also utilized by physical therapists to assist airway clearance and exercise. NIV has been proven to decrease fatigue during airway clearance and reduce oxygen desaturation during exercise. However, it is unclear why and how NIV is implemented during physical therapy treatment in this population.

**What this paper contributes to our knowledge**

NIV was used by physical therapists with low numbers of patients, primarily in adults and people with more severe lung disease. It was more commonly used to assist airway clearance. Physical therapists were initiating NIV and use was relatively aligned with clinical guidelines. Australian physical therapists appear to use conservative pressure settings, reporting improved tolerance of treatments with minimal complications.

